# Quantifying the risk of measles importation and spread in the United States in 2024

**DOI:** 10.1101/2024.06.06.24308559

**Authors:** Subekshya Bidari, Haokun Yuan, Wan Yang

## Abstract

Measles outbreaks have increased globally following the COVID-19 pandemic. We combine multiple data sets on global measles incidence, air travels, and vaccinations to assess the risk of travel-related importation and subsequent dissemination of measles in the United States in 2024, and identify months and states with higher measles outbreak risks.

## Introduction

Following measles elimination in 2000, sporadic cases and sometimes larger outbreaks of measles continue to occur in the United States (US)(1), often due to travel-related importations and subsequent spread in under-vaccinated communities (2, 3). Due to immense public health disruptions during the COVID-19 pandemic, there has been a concerning rise in measles cases globally, and substantial increases have occurred, e.g., by 30-fold in 2023 in the European Region (4). This increase in global measles transmission, combined with the declining vaccination rates, heightens the risk of measles importation and spread in the US. To assess the risk of measles outbreaks in the US in 2024, we calculate the probability of measles spread, encompassing both importation and subsequent dissemination at the national and state levels.

## Methods

We estimated the probability of measles importation and subsequent spread given population susceptibility in each US state and nationwide. The risk of measles importation was computed as the probability of having at least 1 primary measles case from outside the US, and the risk of subsequent measles spread was computed as the probability of having at least 1 secondary measles case following any case importation. The statistical computations and data sources are detailed in the Appendix. Briefly, for each state, we computed the probability of importation based on measles prevalence in each source country and inbound air travel volume, and further combined it with state-level Measles, Mumps, and Rubella (MMR) vaccination coverage to compute the probability of subsequent local spread. We combined the state-specific estimates to compute the national risk. The analysis was carried out for each month of 2024, using reported case data for months with available data (January-April 2024 at the time of this study)(5) and cases data in 2023 for months without real-time data.

## Results

At the national level, we estimate higher risk of measles spread during winter and spring (particularly from January through May; Figure 1B). This estimated peak timing likely arises from an increased importation risk during winter when global measles prevalence tends to be higher, and was consistent with historical records (e.g., during 2001-2016 (1)). However, a large outbreak in Chicago (6) led to the largest reported case count in March, as of May 31, 2024 (Figure 1A). As we focus solely on the initial risk of spread following case importation, our estimates did not capture the magnitude of outbreak as indicated by case counts.

**Figure 1:**
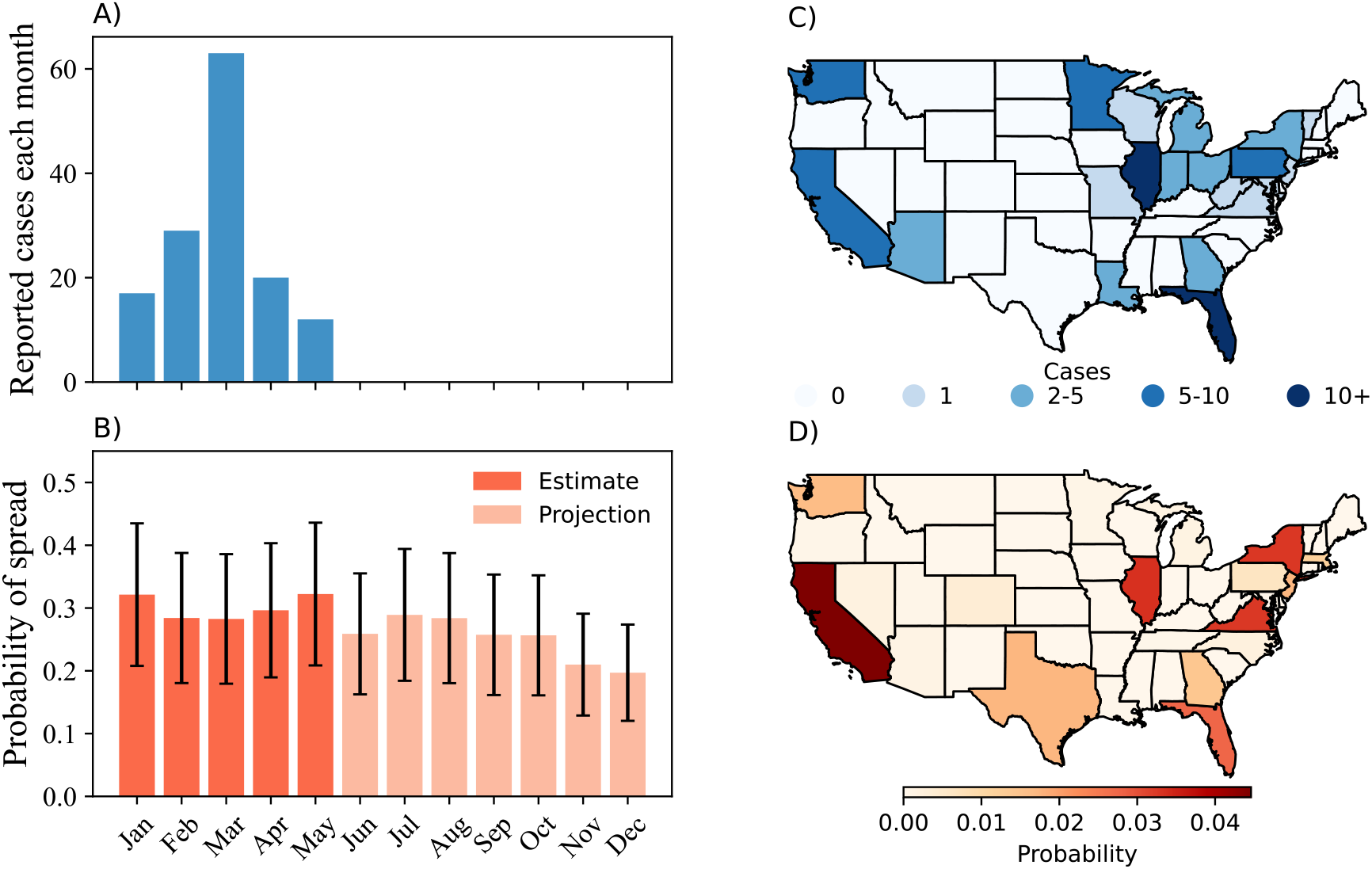
Reported measles case counts and estimated probability of spread in the US. (A) Reported measles case counts nationwide by month; (B) Estimated probability of spread in the US in 2024; (C) Number of cases reported in each US state (as of May 31, 2024); and (D) Estimated probability of measles spread in each state for May 2024.

The estimated spatial pattern by state is generally consistent with the reported case distribution (Figure 1. C vs D). States with higher estimated spread probability tend to be those with major metropolitan centers and airports serving many international locations – particularly, California, New York, Illinois, and Florida, likely due to the higher inbound international travel volume combining with incomplete local vaccination coverage.

We project the risk of measles spread for the remaining months of 2024. While the projected risks vary over time (Figure 2A, for June - August), the states with higher projected spread probability are consistent with those identified for January - May (Figure 1D) and throughout 2024 (Figure 2B-D, for June - August; Figures S1-S2 for all 12 months).

**Figure 2:**
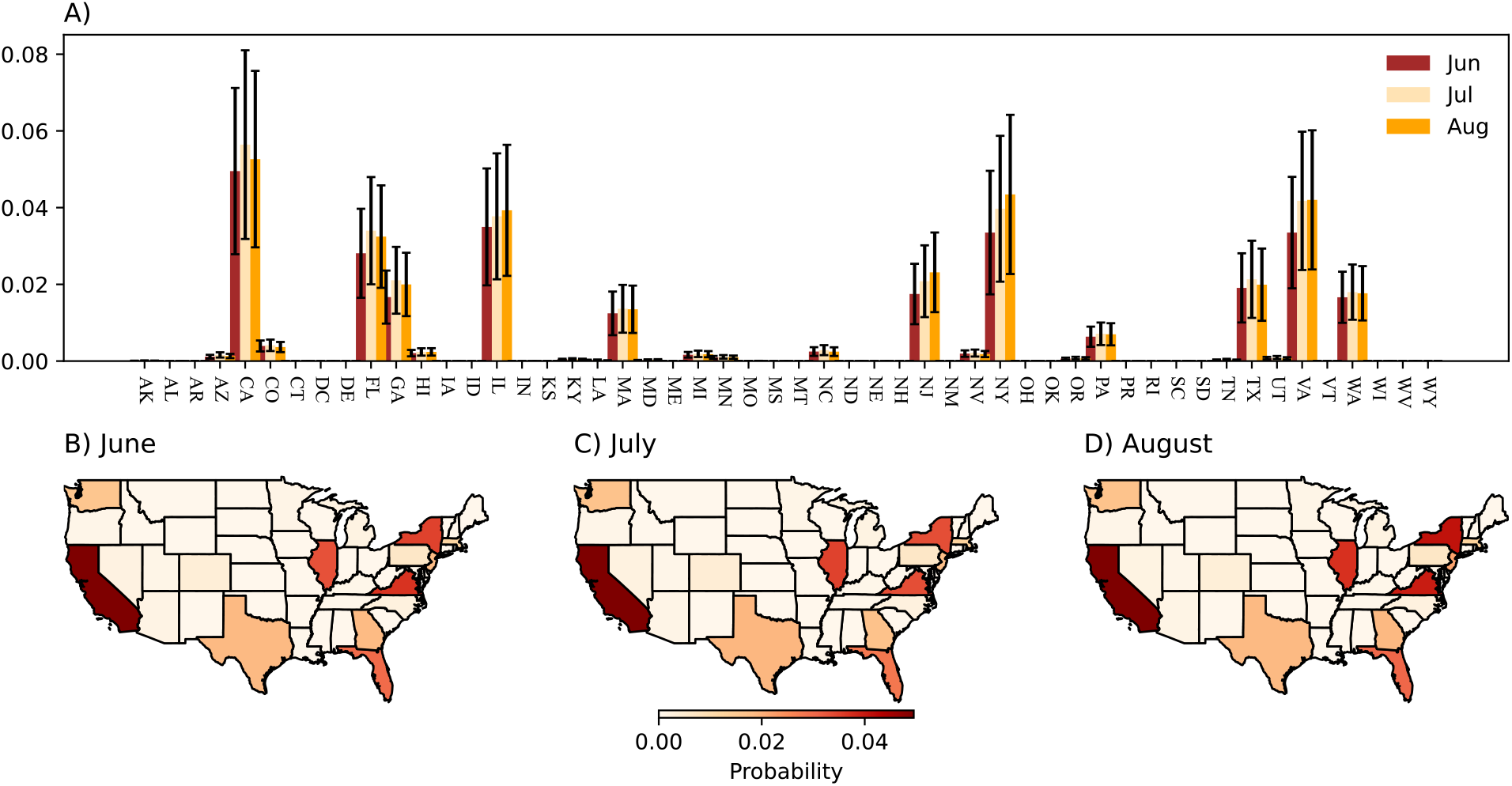
Projected probability of measles spread in the upcoming months in 2024. (A) Colored bars show mean of the projected probability of measles spread from an imported case for the upcoming three months (June, July, August), and vertical lines represent the standard deviation of the estimates. Maps show spatial patterns of projected spread probabilities for (B) June, (C) July, and (D) August. Projections for other months are shown in the Appendix.

## Discussion

An important contribution of our work is identifying states in the US with a higher measles outbreak risk in 2024, given the rise in measles cases globally. These tend to be states with major international airline connections facilitating case importation, in addition to incomplete vaccination coverage. Increased measles surveillance and vaccination efforts in these states could significantly mitigate the risk of measles transmission locally and across the country.

We recognize two main study limitations. First, the travel data only include air travels via direct flights, which could lead to underestimation of risk due to missing sources including those from land travels and countries lacking direct flight service to the US (particularly, those with endemic measles transmission). Second, the study does not account for measles importation between states, thereby overlooking cases such as the ones reported in Louisiana in individuals returning from out of state trips (7).

In summary, we developed methods to assess the risk of measles outbreaks in the US and estimated the outbreak risk by months for each state and nationwide in 2024, based on comprehensive global measles incidence data, international travels, and local vaccination coverage. The estimated spatial pattern is consistent with observations thus far, and the projected outbreak risk by state could inform public health efforts in the coming months.

## Data Availability

All data produced are available online as detailed in the manuscript.

## Acknowledgement

This study was supported by the National Institute of Allergy and Infectious Diseases (grant number: R01AI145883).

## Appendix

### Data

#### Measles data

Global measles case data were compiled from the World Health Organization (WHO) [1]. The most recent monthly data available at the time of analysis was April 2024, and data for the first four months of 2024 were provisional. We used linear interpolation to estimate the number of measles cases for the countries that did not report measles cases for the first four months of 2024. Specifically, we fit country-level linear regression models to measles cases from 2023 and use the slope as an indicator of the measles trend for each country for the months with missing data. A positive (negative) slope signifies increasing (decreasing) trend, meaning measles cases are higher (lower) than the month before. If the 95% confidence interval (CI) of the slope does not include zero (e.g., significant at the α = 0.05 level), we impute missing value using the linear model; and if the 95% CI includes zero but the 68% CI does not, we divide the slope by half and use the linear model for imputation. Otherwise, we assign the average of the monthly measles cases from the previous year to the missing value.

The measles case numbers for US states were compiled from Centers for Disease Control and Prevention (CDC), county level public health organization, and local news reports (summarized in Table S1).

**Table S1:** Measles cases reported in US states in the first five months of 2024.

| State | January | February | March | April | May | Source |
| --- | --- | --- | --- | --- | --- | --- |
| Arizona | 0 | 3 | 2 | 0 | 0 | [2, 3] |
| California | 0 | 2 | 4 | 0 | 3 | [4] |
| Florida | 0 | 10 | 0 | 1 | 0 | [5] |
| Georgia | 1 | 1 | 0 | 1 | 0 | [6, 7, 8] |
| Indiana | 0 | 1 | 0 | 1 | 0 | [9] |
| Louisiana | 0 | 2 | 0 | 0 | 0 | [10] |
| Maryland | 0 | 1 | 0 | 0 | 0 | [11] |
| Michigan | 0 | 1 | 2 | 0 | 0 | [12, 13, 14] |
| Minnesota | 0 | 3 | 0 | 3 | 3 | [15] |
| Missouri | 1 | 0 | 0 | 0 | 0 | [16] |
| New Jersey | 1 | 0 | 0 | 0 | 0 | [17] |
| New York | 0 | 2 | 1 | 0 | 0 | [18, 19] |
| Ohio | 1 | 2 | 1 | 0 | 0 | [20, 21, 22, 23] |
| Pennsylvania | 9 | 0 | 0 | 0 | 1 | [24, 25] |
| Virginia | 1 | 0 | 0 | 0 | 0 | [26] |
| Washington | 3 | 1 | 0 | 0 | 2 | [27, 28, 29, 30] |
| Illinois | 0 | 0 | 53 | 11 | 3 | [31] |
| Vermont | 0 | 0 | 0 | 0 | 1 | [32] |
| West Virginia | 0 | 0 | 0 | 0 | 1 | [33] |
| Wisconsin | 0 | 0 | 0 | 0 | 1 | [34] |

#### Air travel data

Data on air travel volume were obtained from the Bureau of Transportation Statistics (BTS) [35]. The BTS releases data on all flights operated by both US and foreign air carriers with either origin or destination located within the boundaries of the US. We used the monthly travel data with destination in any US state for the year 2023 for the analysis.

#### Population data

Population data for the origin countries were obtained from the World Bank database [36]. We used population data for 2022, which was the most recent year with available data. The population data for US states were obtained from the US Census Bureau [37] and the estimates for 2023 were used.

#### Vaccination rates

We calculate population level vaccination coverage for US states using the data on Vaccination Coverage and Exemptions among Kindergartners provided by the National Center for Immunization and Respiratory Diseases [38]. Combining the Measles, Mumps, and Rubella (MMR) vaccination rates dating back to 2009-10 and the fraction of population in different age groups [37], we obtain a population level estimate of MMR vaccination rate for each US state.

### Methods

We compute the probability of importing at least one case of measles and probability of measles spread (secondary transmission beyond the imported case) for each state in the US using two main factors - air travel volumes from countries currently experiencing measles outbreaks and the population level MMR vaccination rates in each state.

- Denote the locations (other countries) with direct air travel destinations in the US states as origin O and the list of US states as D. We convert monthly measles incidence at the origin, *M*_*o*_ into prevalence of infectious people *I*_*o*_ (i.e., the average number of infectious people present in the origin population, during month m)

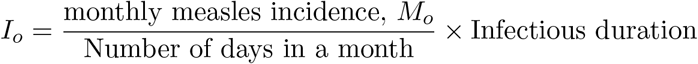
- Obtain fraction of infectious people at the origin *p*_*o*_ (i.e., the average probability of a traveler from the origin country O being infectious)

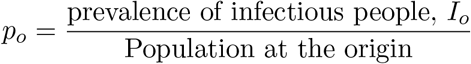
- Denote *A*_*ij*,*m*_ as the volume of air travel between origin *i* ∈ *O* and destination *j* ∈ *D* in month *m*. The pair-wise probability of importation for a origin-destination pair *ij* is computed as

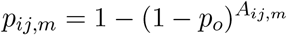
- The aggregate probability of importation for each destination state *j* ∈ *D* for month m is then calculated as

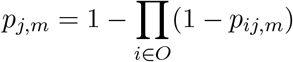
- Denote *v*_*j*_, *HH*_*SAR*_ and *O*_*SAR*_ as the population level vaccination rate for each destination state, secondary attack rates for measles within household and outside household, respectively. The pair-wise probability of measles spread from *i* ∈ *O* to a state *j* ∈ *D* is estimated as

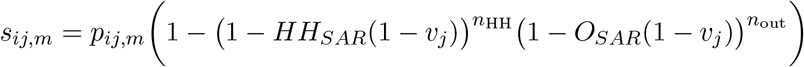
- The aggregate probability of spread for each state is calculated as

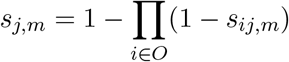

Due to a lack of data on measles secondary attack rates outside the household, we estimate measles secondary attack rate for contacts outside the household using the ratio of household and non-household secondary attack rates reported for Covid-19 in previous studies [39, 40, 41]. To account for the uncertainty in model variables, we run the analysis using a range of plausible values and report the mean and standard error for the analysis. We assume infectious period duration of 3 – 7 days, average household contact *n*_HH_ ∈ [2, 3], average contact outside the household *n*_out_ ∈ [10, 20, 30, 40], *HH*_*SAR*_ = 0.9, and *O*_*SAR*_ ∈ [0.18, 0.27, 0.36, 0.45].

## Supplemental Figures

**Figure S1:**
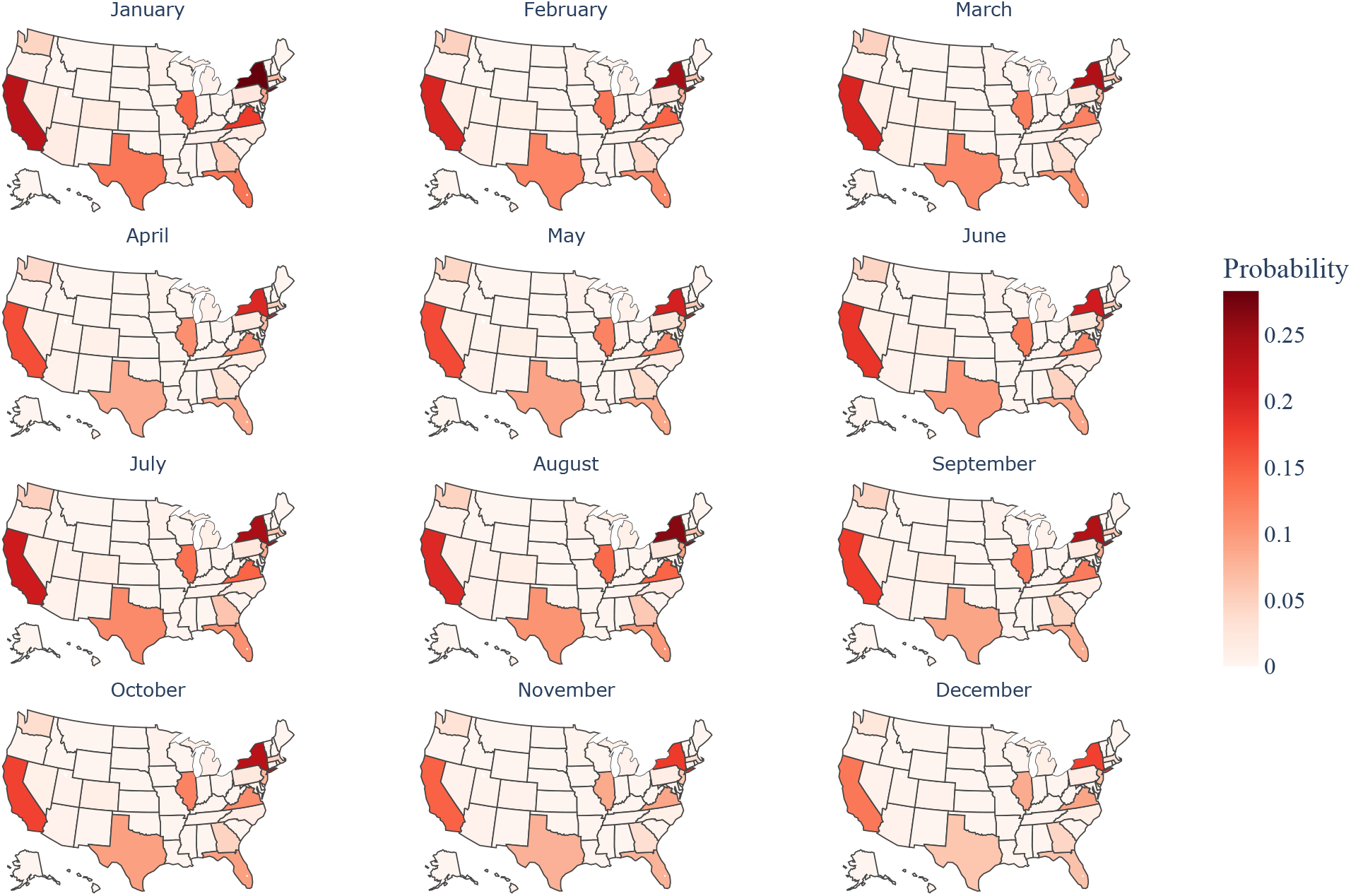
Estimated probability of measles importation in 2024

**Figure S2:**
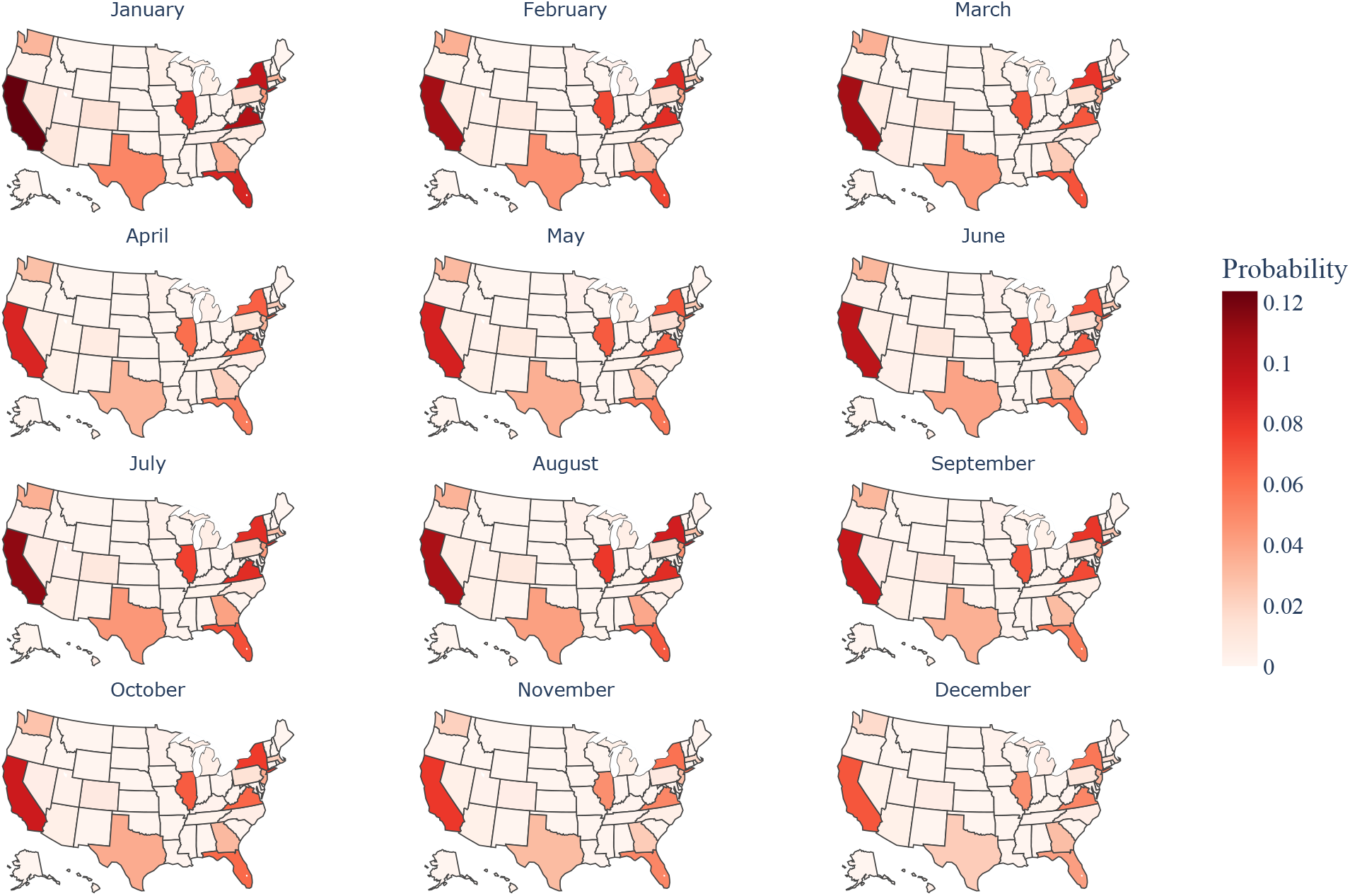
Estimated probability of measles spread in 2024

